# The NLRP3 inflammasome as a key pathway in the affective and chronic fatigue symptoms of Long COVID

**DOI:** 10.1101/2025.09.07.25335261

**Authors:** Yingqian Zhang, Hussein Kadhem Al-Hakeim, Hawraa Kadhem Al-Jassas, Michael Maes

**Affiliations:** Sichuan Provincial Center for Mental Health, Sichuan Provincial People’s Hospital, School of Medicine, University of Electronic Science and Technology of China, Chengdu 610072, China; Key Laboratory of Psychosomatic Medicine, Chinese Academy of Medical Sciences, Chengdu, 610072, China; Department of Chemistry, Faculty of Sciences, University of Kufa, Iraq; Department of Pharmaceutical Chemistry, Faculty of Pharmacy, University of Kufa, Iraq; Department of Psychiatry, Faculty of Medicine, Chulalongkorn University, Bangkok, Thailand; Department of Psychiatry, Medical University of Plovdiv, Plovdiv, Bulgaria; Research Institute, Medical University of Plovdiv, Plovdiv, Bulgaria; Research and Innovation Program for the Development of MU - PLOVDIV (SRIPD-MUP), Creation of a network of research higher schools, National Plan for Recovery and Sustainability, European Union – NextGenerationEU, Medical University of Plovdiv, Plovdiv, Bulgaria, Europe; Kyung Hee University, Seoul, Dongdaemun-gu, South Korea

**Keywords:** Long COVID, NLRP3, chronic fatigue, inflammation, depression, biomarkers

## Abstract

The neuropsychiatric and somatic manifestations (physio-affective phenome) of Long COVID are substantially predicted by elevated peak body temperature (PBT) and diminished oxygen saturation (SpO_2_) during the acute infectious stage. The latter is linked to the immune pathophysiology of Long COVID involving activation of the immune-inflammatory response system (IRS) and the NLRP3 inflammasome. Nevertheless, there is a lack of data indicating whether NLRP3 and its components are implicated in the physio-affective phenome of Long COVID.

We enrolled 161 Long COVID patients 6 to 9 months after the acute phase and divided them into two groups based on the baseline PBT and SpO_2_ levels, namely mild and severe acute COVID-19. We assessed serum NLRP3, caspase-1, C-reactive protein (CRP), interleukin (IL)-18, IL-1β, IL-10, fibronectin, and Gasdermin D (GSDMD) during Long COVID.

All of the aforementioned indicators (with the exception of IL-10) were substantially higher in Long COVID patients who had previously experienced severe COVID-19 than in those who had mild acute COVID-19. The physio-affective phenome of Long COVID and the severity of the acute COVID-19 were significantly correlated with these IRS biomarkers, with the exception of fibronectin. The variance in the overall severity of Long COVID is accounted for (49.5%) by the combined influence of fibronectin, IL-10, SpO_2_, and PBT.

In conclusion, the severity of Long COVID is strongly associated with IRS and NRLP3 activation and the severity of the inflammatory response during the acute infectious phase. NLRP3 activation is a drug target to treat Long COVID.

## Introduction

Coronavirus disease-2019 (COVID-19) infection ranks among the most prevalent contagious diseases in 2022. While COVID-19 patients can recover, certain individuals experience ongoing symptoms or endure a new condition following COVID-19, which is termed long COVID (Venkatesan, 2021; 2024). This syndrome encompasses a range of enduring neuropsychiatric illnesses, including emotional symptoms (anxiety, depression), and many physio-somatic symptoms, including chronic fatigue syndrome (CFS), fibromyalgia, cognitive deficits, respiratory issues, and dyspnea (Kubota *et al*., 2023; Greenhalgh *et al*., 2024). One latent construct underpins these neuropsychiatric and chronic fatigue symptoms, and it is named the “physio-affective phenome” (Al-Jassas *et al*., 2022). The severity of the phenome can be calculated as the first factor derived from the neuropsychiatric rating scales, including the Fibro-Fatigue (FF) scale and the Hamilton Anxiety (HAMA) and Depression (HAMD) Rating Scales (Al-Hakeim *et al*., 2022; Al-Hadrawi *et al*., 2023; Al-Hakeim *et al*., 2023).

Earlier studies suggest that changes in peak body temperature (PBT) (increased) coupled with oxygen saturation (SpO_2_) (decreased) during acute SARS-CoV-2 infection account for much of the variance in Long COVID’s overall severity of illness (labeled the physio-affective phenome) (Al-Jassas *et al*., 2022). This indicates that activation of the immune-inflammatory response system (IRS) during acute SARS-CoV-2 infection significantly influences the development of Long COVID(Almulla *et al*., 2022; Al-Hadrawi *et al*., 2023). Since lowered SpO_2_ and elevated PBT indicate the severity of illness and inflammation, we created a new indicator, z PBT – z SpO_2_, to facilitate data analysis.

The IRS pathophysiology of the acute phase of COVID-19 is related to increased nucleotide-binding domain leucine-rich repeat and pyrin domain-containing receptor 3 (NLRP3) inflammasome activity in the peripheral blood and tissues of postmortem patients (Bryant, 2021; Rodrigues *et al*., 2021a). NLRP3 is involved in inflammatory and autoinflammatory illnesses and is an essential part of the innate immune system (Karen V Swason *et al*., 2019). The NLRP3 protein is primarily made up of three components: a central nucleotide-binding domain (NACHT), a pyrin domain (PYD), and a leucine-rich repeat (LRR) domain (Sharif *et al*., 2019).

During NLRP3 priming, Toll-like receptors identify PAMPs and DAMPs (pathogen-and damage-associated molecular patterns, respectively), including fibronectin, which trigger nuclear factor kappa B (NF-κB) signaling. Then, NF-κB induces NLRP3 transcription and pro-IL-1β, pro-caspase-1, and pro-IL-18 (Zhao *et al*., 2021; He *et al*., 2024). This activation step is primarily triggered by ATP, viral RNA, etc., causing the formation of the NLRP3 and pro-caspase-1 complex(He *et al*., 2016). Several molecular and cellular events participate in the activation step, such as lysosomal rupture, increased reactive oxygen species (ROS), and mitochondrial aberrations (Zhao *et al*., 2021; Rodrigues & Zamboni, 2023).

Long COVID is correlated with IRS activation and is accompanied by increased C-reactive protein (CRP) levels compared to controls, suggesting a mild inflammatory response(Al-Hakeim *et al*., 2022). A review summarized that Long COVID is coupled with inflammation, including reduced serum albumin, elevated neutrophils, and lipid abnormalities (Mehandru & Merad, 2022). A recent meta-analysis showed an increased IRS to compensatory immunoregulatory system (CIRS) ratio and increased T helper (Th)1, Th17, and immune-associated neurotoxicity in Long COVID (Almulla *et al*., 2024).

The NLRP3 inflammasome, part of the innate immune system, is necessary for defending against viruses, while abnormal activation may result in tissue damage and neuropsychiatric symptoms (van den Berg & te Velde, 2020; Maes *et al*., 2025). In addition, NLRP3 activation might play a vital role in the pathophysiology of acute COVID-19 (Tisoncik *et al*., 2012; Lin *et al*., 2019). Ribeiro et al. reviewed that the IRS response to infection is triggered by P2X7 receptor engagement, leading to NLRP3 activation, thereby causing COVID-19-associated neuropsychiatric conditions (Ribeiro *et al*., 2021). Current evidence shows that increased NLRP3 inflammasome is related to depression or anxiety stemming from Long COVID(Ribeiro *et al*., 2021). However, there is an absence of data showing that NLRP3 and IL-1β, IL-10, IL-18, etc., are part of the physio-affective phenome of Long COVID.

Hence, this study was conducted to examine: 1) the difference in physio-affective symptoms and IRS biomarkers during Long COVID in individuals who had previously suffered from severe versus mild SARS-CoV-2 infection; and 2) the association between IRS biomarkers, including NLRP3, and Long COVID’s severity. The hypotheses specifically proposed that NLRP3 activation is associated with Long COVID’s physio-affective phenome and is predicted by lowered SpO_2_ or increased PBT.

### Subjects and Methods Participants

This research examined disparities between male Long COVID participants categorized into two groups based on the severity of inflammatory markers during the acute infection, specifically PBT and SpO_2_ values. The Long COVID patients were categorized into two groups based on a composite score derived from the values ( z PBT – z SpO_2_), referred to as the IPS index, which stands for inflammation, <u>P</u>BT, and <u>S</u>pO_2_: those with a low IPS index (n = 81, mild acute COVID-19) and those with a high IPS index (n = 80, severe acute COVID-19). The period for enrolling participants in this study began in June 2024 and ended in September 2024. All persons identified with Long COVID had previously undergone treatment for acute COVID-19 infection at one of the designated quarantine hospitals in Kerbala City, Iraq. The hospitals comprise Al-Sadr Medical City and Al-Hakeem General Hospital in Najaf City, Iraq.

The diagnosis of acute infection was established by senior physicians and virologists: a) the presence of infectious symptoms such as fever, cough, respiratory difficulties, and anosmia; b) positive IgM antibodies against SARS-CoV-2; and c) affirmative results from reverse transcription real-time polymerase chain reaction (RT-PCR) testing. Long COVID was diagnosed 6 to 9 months post-acute infection by the same senior physicians and virologists. The diagnosis was established according to the WHO Long COVID case definition(Venkatesan, 2021), which encompasses i) continued or new-onset symptoms beyond the initial phase of the illness; ii) symptoms persist for a minimum of 2 months and are evident 3-4 months following the infection; iii) at least two symptoms that disrupt daily activities, including loss of smell or taste, persistent cough, chest pain, fever, headache, affective symptoms, cognitive impairment, fatigue, or difficulty speaking; and iv) a diagnosis of SARS-CoV-2 infection. The results of the RT-PCR tests indicated a negative outcome for SARS-CoV-2 during the Long COVID period.

The study excluded individuals with a) systemic immune disorders, such as chronic fatigue syndrome, chronic obstructive pulmonary disease, type 1 diabetes mellitus, inflammatory bowel disease, scleroderma, rheumatoid arthritis, psoriasis, and other infectious disorders; b) individuals with a history of psychiatric disorders, including bipolar disorder, major depressive disorder, schizophrenia, generalized anxiety disorder, dysthymia, post-traumatic stress disorder, substance use disorders, and autism spectrum disorder; c) neurologic disease, including stroke, multiple sclerosis, and Alzheimer’s and Parkinson’s disease; d) liver and kidney disease.

Upon acquiring thorough information, all participants, along with their legal guardians, provided informed consent prior to the initiation of the study. The study received approval from the Kerbala Health Directorate-Training and Human Development Center, as well as the institutional ethics committee of the University of Kufa, as indicated by the documentation bearing the reference numbers 4316-2024/1/29.

### Clinical measurements

A senior pulmonologist conducted an evaluation and gathered sociodemographic and clinical data from patients experiencing Long COVID, at least six months following the acute infectious phase. This was achieved through semi-structured interviews and a thorough examination of patient records, which included pertinent information from the acute phase of their illness. Levels of IgM antibodies against SARS-CoV-2, results from RT-PCR, PBT, SpO_2_ (%), and the symptoms associated with acute COVID-19 infection during the acute phase of the illness were meticulously extracted from the patient records. During the acute phase of infection, a paramedical specialist conducted a precise assessment of SpO_2_ levels utilizing a state-of-the-art electronic oximeter manufactured by Shenzhen Jumper Medical Equipment Co., Ltd. The measurement of PBT was conducted utilizing a digital oral thermometer, which was strategically placed beneath the tongue until an audible beep indicated the completion of the reading.

In light of the PBT and SpO_2_ values acquired during the acute infectious phase, we have developed a novel indicator that integrates the decrease in SpO_2_ alongside the increase in body temperature. This is quantified as the z transformation of PBT (z PBT) subtracted by z SpO_2_ (Al-Hakeim et al., 2023). By employing the median of this composite value (labeled IPS index-from inflammation, <u>P</u>BT, and <u>S</u>pO_2_), we systematically classify the individuals experiencing Long COVID into two distinct groups: those exhibiting lower IPS values (less than 0.41) and those demonstrating higher IPS composite scores (greater than 0.41). The latter cohort represents a specific subgroup of individuals diagnosed with Long COVID, demonstrating signs of inflammation, as evidenced by elevated PBT and reduced SpO_2_ levels during the acute phase of the illness.

During the Long COVID phase, a senior psychiatrist employed a semi-structured interview methodology to assess sociodemographic and clinical data. On the same day, this physician conducted an evaluation using the Hamilton Depression Rating Scale (HAMD) to assess the severity of depression, the Hamilton Anxiety Rating Scale (HAMA) to measure the severity of anxiety, and the Fibro-Fatigue (FF) scale to quantify the severity of symptoms associated with chronic fatigue syndrome (CFS).

Considering that both the HAMD and the HAMA encompass a variety of physio-somatic symptoms, which may obfuscate the interpretation of depressive and anxiety symptoms within the framework of a medical condition, we have computed the aggregate of the primary affective depressive symptoms from the HAMD and the principal affective symptoms from the HAMA, deliberately excluding the physio-somatic symptoms identified in those scales. The principal symptoms of “pure” HAMD were quantified by aggregating sad mood, feelings of guilt, suicidal ideation, and lack of interest, collectively referred to as “pure depression.” The principal symptoms of anxiety as measured by the HAMA were derived from the cumulative assessment of anxious mood, tension, fears, and anxiety-related behaviors observed during the interview, collectively termed “pure anxiety.” In order to conduct a precise assessment of the physio-somatic symptoms associated with the FF, we have deliberately excluded any symptoms that pertain to sadness, cognitive disturbances, insomnia, and irritability. Consequently, the symptoms associated with “pure” CFS symptoms were quantified by aggregating muscular pain, muscle tension, fatigue, autonomic symptoms, headaches, gastrointestinal symptoms, and flu-like malaise. The overall severity of Long COVID (OSOLC) was calculated as a composite score utilizing z units, which amalgamated z pure HAMD, z pure HAMA, and z pure FF. The calculation for body mass index (BMI) involves dividing the mass in kilograms by the square of the height measured in meters.

### Assays

After an overnight fast, about 5 mL of venous blood was drawn using disposable syringes between 7:00 and 9:00 am. The blood was collected into sterile serum tubes, then centrifuged at 3500 rpm for 10 minutes at room temperature. The serum was aliquoted for several Eppendorf tubes, stored at -80 until they were thawed for assaying. The serum levels of NLRP3, CRP, fibronectin, GSDMD, caspase-1, IL-1β, IL-10, and IL-18 were measured by ELISA kits obtained from Nanjing Pars Biochem Co., Ltd. (Nanjing, China). The intra-assay and inter-assay analytical coefficients of variation (CVs) were <10% and 12%, respectively.

### Statistics

The interrelationships among scale variables were examined through the application of Spearman’s rank-order correlation coefficients, Pearson’s product-moment correlation, and partial correlation coefficients. A one-way analysis of variance was employed to compare scale variables among different groups, whereas chi-square tests were utilized to evaluate the relationships between categorical variables. Multiple comparisons or correlations were adjusted for false discovery rate (FDR) to mitigate the risk of type I errors. A comprehensive multiple regression analysis was performed to ascertain the principal biomarkers that predict the neuropsychiatric symptom domains associated with Long COVID. This analysis employed both manual and stepwise automatic methods, utilizing a *P*-to-entry threshold of 0.05 and a *P*-to-remove threshold of 0.06. We calculated the standardized beta coefficients for each significant explanatory variable by employing t statistics accompanied by accurate p-values, in addition to the model’s F statistics and the total variance explained (R^2^), which functioned as the measure of effect size. Moreover, the evaluation of the analysis for homoscedasticity was conducted employing the White and Breusch-Pagan tests, in addition to examining collinearity concerns through the use of VIF and tolerance metrics.

## Results

### Differences between Long COVID subgroups

The sociodemographic and clinical characteristics of the Long COVID patients, categorized according to the severity of the acute infectious phase, are presented in **Table 1**. The initial three rows present the variables employed to dichotomize the Long COVID cohort into subgroups, specifically according to diminished SpO_2_ levels and elevated PBT values. The two groups exhibited no significant disparities in terms of age, BMI, employment status, marital status, living arrangements, or smoking behaviors. The scores on all rating scales acquired during the Long COVID phase were markedly elevated in patients exhibiting severe inflammation during the acute phase of COVID-19, in comparison to those with lesser degrees of inflammation. This suggests that elevated levels of inflammation during the acute phase of COVID-19 are predictive of increased affective and physio-somatic symptoms in the subsequent months.

**Table 1.**
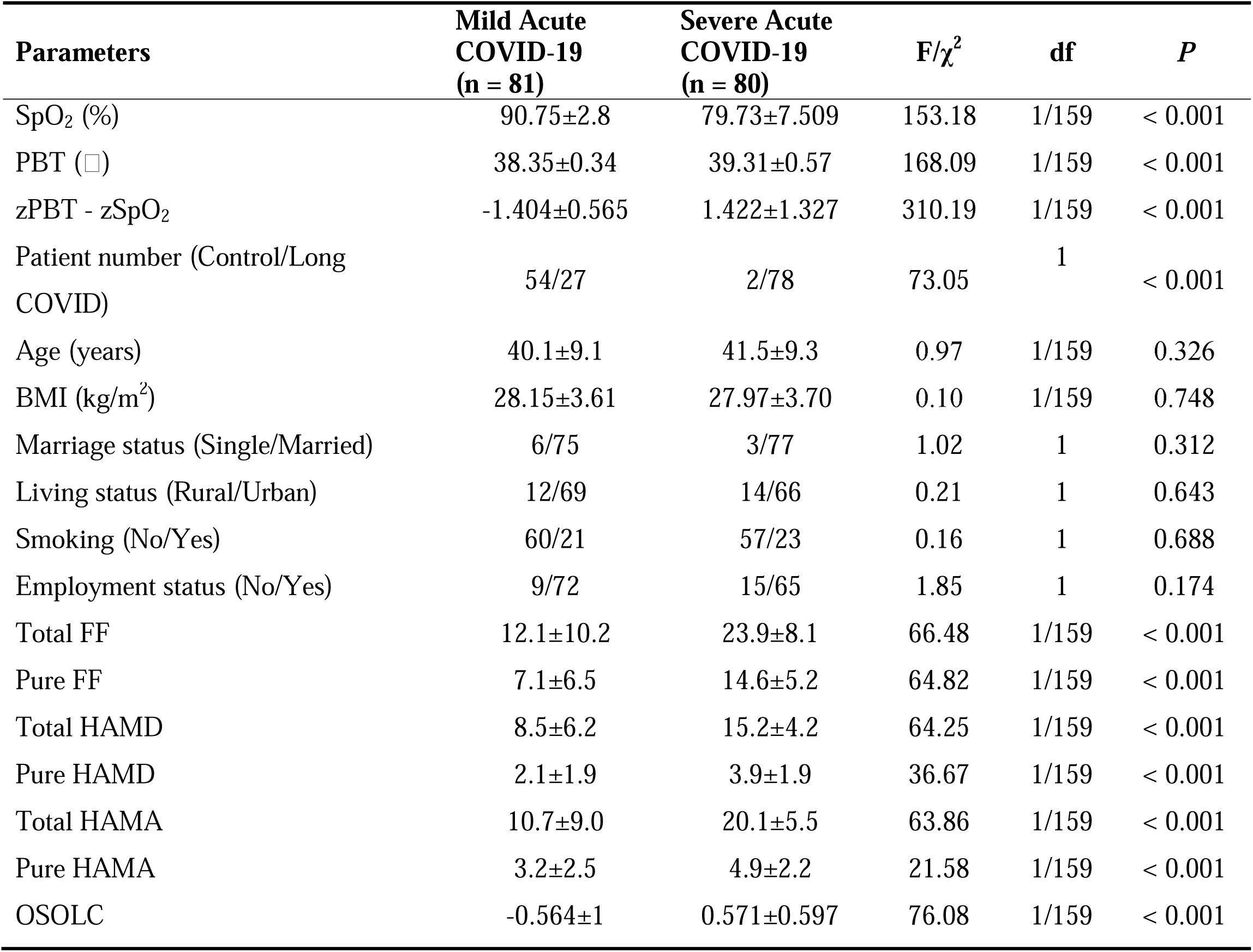
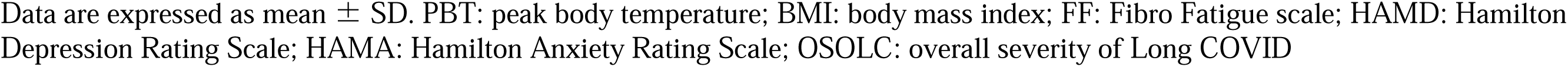
Demographic and clinical data of Long COVID patients divided into those with mild versus severe acute COVID-19 infection.

| Parameters | Mild Acute<br>COVID-19<br>(n = 81) | Severe Acute<br>COVID-19<br>(n = 80) | F/$\chi^2$ | df | P |
| --- | --- | --- | --- | --- | --- |
| SpO<sub>2</sub> (%) | 90.75±2.8 | 79.73±7.509 | 153.18 | 1/159 | < 0.001 |
| PBT (□) | 38.35±0.34 | 39.31±0.57 | 168.09 | 1/159 | < 0.001 |
| zPBT - zSpO<sub>2</sub> | -1.404±0.565 | 1.422±1.327 | 310.19 | 1/159 | < 0.001 |
| Patient number (Control/Long<br>COVID) | 54/27 | 2/78 | 73.05 | 1 | < 0.001 |
| Age (years) | 40.1±9.1 | 41.5±9.3 | 0.97 | 1/159 | 0.326 |
| BMI (kg/m<sup>2</sup>) | 28.15±3.61 | 27.97±3.70 | 0.10 | 1/159 | 0.748 |
| Marriage status (Single/Married) | 6/75 | 3/77 | 1.02 | 1 | 0.312 |
| Living status (Rural/Urban) | 12/69 | 14/66 | 0.21 | 1 | 0.643 |
| Smoking (No/Yes) | 60/21 | 57/23 | 0.16 | 1 | 0.688 |
| Employment status (No/Yes) | 9/72 | 15/65 | 1.85 | 1 | 0.174 |
| Total FF | 12.1±10.2 | 23.9±8.1 | 66.48 | 1/159 | < 0.001 |
| Pure FF | 7.1±6.5 | 14.6±5.2 | 64.82 | 1/159 | < 0.001 |
| Total HAMD | 8.5±6.2 | 15.2±4.2 | 64.25 | 1/159 | < 0.001 |
| Pure HAMD | 2.1±1.9 | 3.9±1.9 | 36.67 | 1/159 | < 0.001 |
| Total HAMA | 10.7±9.0 | 20.1±5.5 | 63.86 | 1/159 | < 0.001 |
| Pure HAMA | 3.2±2.5 | 4.9±2.2 | 21.58 | 1/159 | < 0.001 |
| OSOLC | -0.564±1 | 0.571±0.597 | 76.08 | 1/159 | < 0.001 |

Data are expressed as mean ± SD. PBT: peak body temperature; BMI: body mass index; FF: Fibro Fatigue scale; HAMD: Hamilton Depression Rating Scale; HAMA: Hamilton Anxiety Rating Scale; OSOLC: overall severity of Long COVID

**Table 2** presents the biomarkers associated with Long COVID across the two subgroups examined. Our findings indicate that NLRP3, caspase-1, GSDMD, fibronectin, IL-18, and IL-1β were significantly elevated in Long COVID patients who exhibited heightened inflammatory markers during the acute infectious phase, in contrast to those with a lower IPS score. Notably, IL-10 did not show a significant increase in this context. Despite the application of FDR *P* correction, the observed differences remained statistically significant; for instance, NLRP3 exhibited a *P* value of 0.029, while caspase-1 demonstrated a *P* value of 0.021. Consequently, the IPS score observed during the acute phase serves as a significant predictor of more severe affective and physio-somatic symptoms.

**Table 2.** Differences in biomarkers of Long COVID between subjects with mild versus severe acute COVID-19 infection.

| **Biomarkers of Long COVID** | **Mild Acute**<br>**COVID-19**<br>**(n = 81)** | **Severe Acute**<br>**COVID-19**<br>**(n = 80)** | **F** | **P** |
| --- | --- | --- | --- | --- |
| NLRP3 (pg/mL) | 31.41(1.44) | 36.03(1.45) | 5.09 | 0.026 |
| Caspase-1 (pg/mL) | 7.66(1.10) | 11.07(1.11) | 5.86 | 0.017 |
| GSDMD (pg/mL) | 40.19(2.58) | 56.95(2.60) | 20.91 | < 0.001 |
| Fibronectin (ng/mL)* | 458.39(23.27) | 556.44(23.42) | 8.04 | 0.005 |
| IL-18 (pg/mL) | 127.62(11.33) | 204.616(11.41) | 22.85 | < 0.001 |
| IL-1β (pg/mL)* | 6.36(0.81) | 11.628(0.81) | 34.54 | < 0.001 |
| IL-10 (pg/mL) | 6.63(0.54) | 8.02(0.55) | 3.24 | 0.074 |

Data are expressed as estimated marginal mean (SE) values obtained by GLM analysis (df=1/154) after covarying for age, sex, education, and smoking. NLRP3: nucleotide-binding domain leucine-rich repeat and pyrin domain containing receptor 3; GSDMD: Gasdermin D; IL: interleukin; \* Processed in log transformation

### Associations between severity of illness and biomarkers

**Table 3** presents the Pearson correlation between the z PBT – z SpO_2_ composite score during the acute phase of illness and the overall severity of Long COVID (OSOLC), alongside the associated biomarkers of Long COVID. Our findings indicate that the z PBT – z SpO_2_ composite score exhibited a significant association with all biomarkers, with the exception of fibronectin. All these differences continued to be significant following the FDR *P* correction, as exemplified by caspase-1 at *P* = 0.0495. Following the application of FDR *P* corrections, a significant correlation was observed between the OSOLC and all biomarkers.

**Table 3.** Associations between the severity of the acute SARS-CoV-2 infectious phase and the biomarkers and overall severity of Long COVID (OSOLC)

|  | **z PBT – z SPO<sub>2</sub>** | **OSOLC** |
| --- | --- | --- |
| OSOLC | 0.508 (< 0.001) | - |
| NLRP3 | 0.185 (0.019) | 0.32 (< 0.001) |
| CRP | 0.444 (< 0.001) | 0.427 (< 0.001) |
| Caspase-1 | 0.159 (0.044) | 0.310 (< 0.001) |
| GSDMD | 0.226 (0.004) | 0.298 (< 0.001) |
| Fibronectin | 0.139 (0.078) | 0.338 (< 0.001) |
| IL-18 | 0.399 (< 0.001) | 0.409 (< 0.001) |
| IL-1β | 0.313 (< 0.001) | 0.502 (< 0.001) |
| IL-10 | 0.168 (0.033) | 0.395 (< 0.001) |

Data are expressed as estimated marginal mean (SE) values obtained by Pearson's correlation coefficients (n=161). OSOLC: overall severity of Long COVID; NLRP3: nucleotide-binding domain leucine-rich repeat and pyrin domain containing receptor 3; CRP: C-reactive protein; GSDMD: Gasdermin D; IL: interleukin.

**Table 4** delineates the results of multiple regression analyses, wherein clinical rating scale scores serve as the dependent variables. The analysis incorporates all biomarkers associated with Long COVID, both with and without the inclusion of PBT and SpO_2_, evaluated during the acute phase of infection as predictors. Our analysis revealed that 45.7% of the variance in OSOLC can be attributed to the cumulative influences of IL-1β, IL-10, CRP, IL-18, and fibronectin, all of which exhibited positive correlations. **Figure 1** illustrates the partial regression analysis of the composite score in relation to IL-1β levels observed in patients suffering from Long COVID. In the context of our regression analysis, the introduction of PBT and SpO_2_ revealed that 49.5% of the variance is accounted for by SpO_2_, which exhibits a negative correlation, alongside IL-1β, IL-10, fibronectin, and PBT, all of which are positively associated. **Figure 2** illustrates the partial regression analysis of the OSOLC composite in relation to the SpO_2_ levels observed during the acute phase of infection. The variables PBT and SpO_2_ accounted for 28.9% of the variance observed in OSOLC. A significant portion of the variance in the pure FF score can be attributed to IL-1β, IL-10, CRP, IL-18, and fibronectin, all of which exhibit positive correlations. Our findings indicate that as much as 40% of the variance in the pure HAMD score can be accounted for by the levels of IL-10, IL-1β, CRP, and IL-8, all of which exhibit a positive association. The combination of IL-1β and IL-10 accounted for 25.2% of the variance observed in the pure HAMA score.

**Figure 1:**
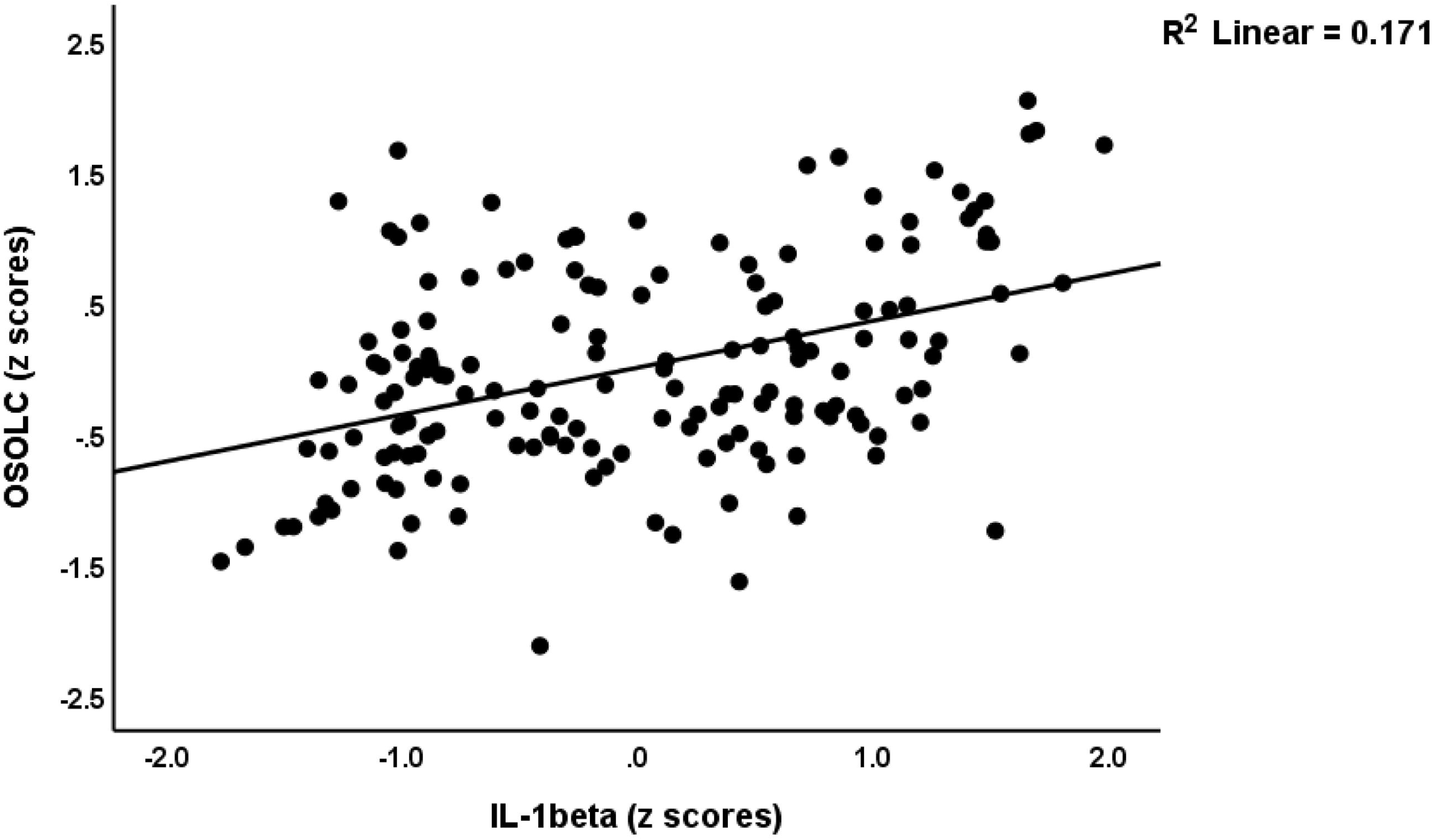
Partial regression of the overall severity of Long COVID on serum IL-1β levels in Long COVID patients (*P* < 0.001).

**Figure 2:**
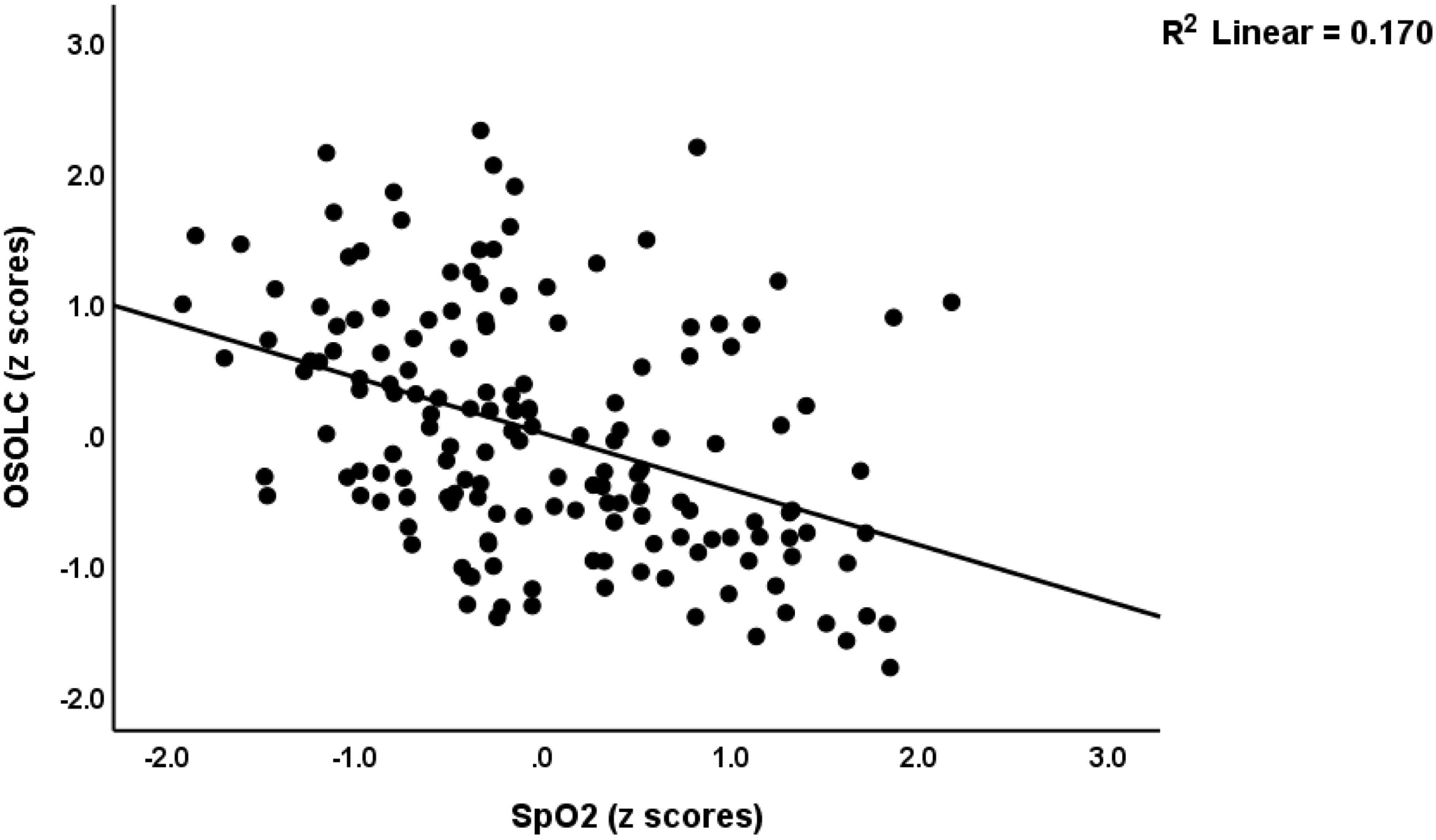
Partial regression of the overall severity of Long COVID (OSOLC) composite score on peripheral blood oxygen saturation (SpO_2_ ) levels during the acute phase of infection (*P* <

**Table 4.**
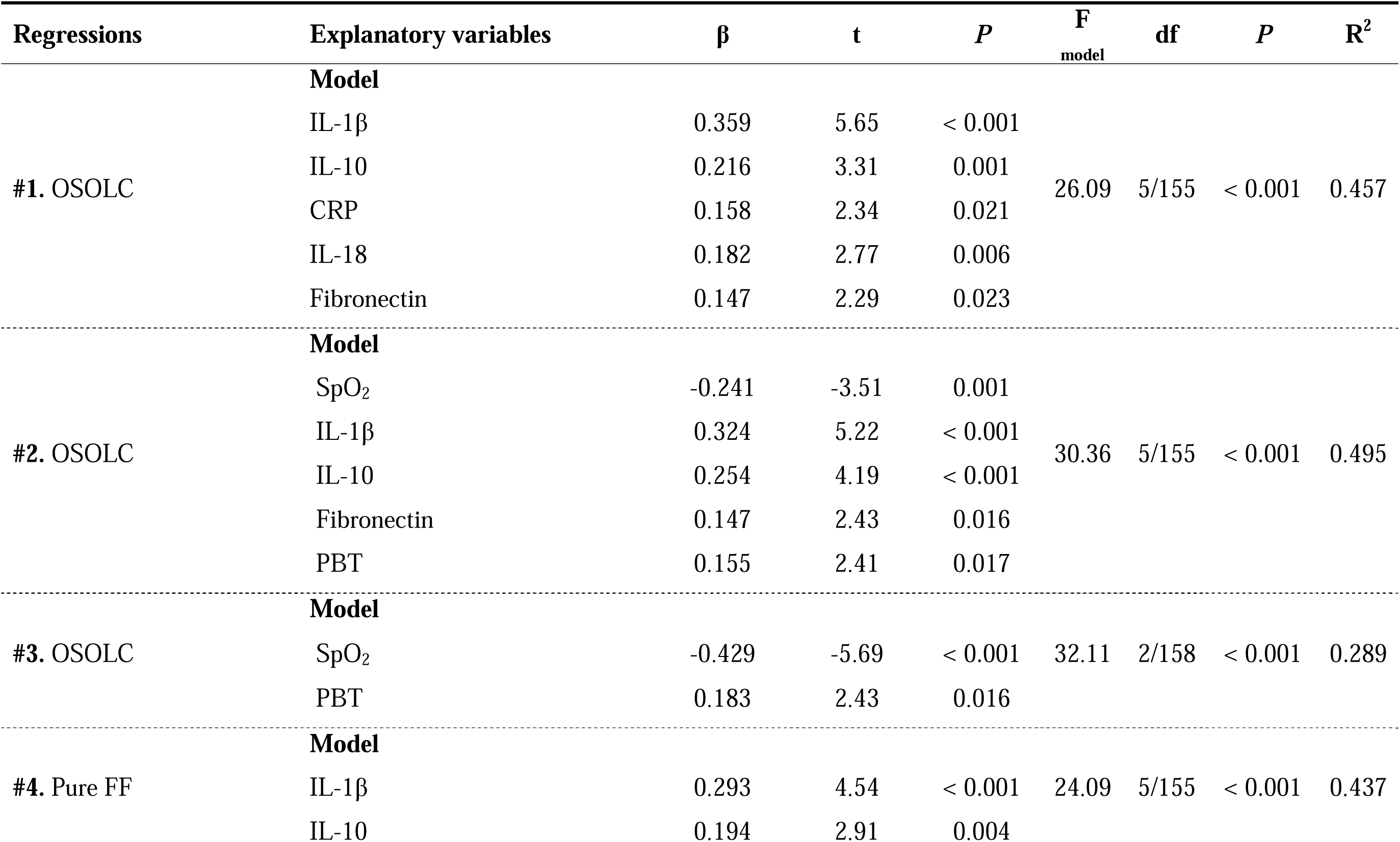
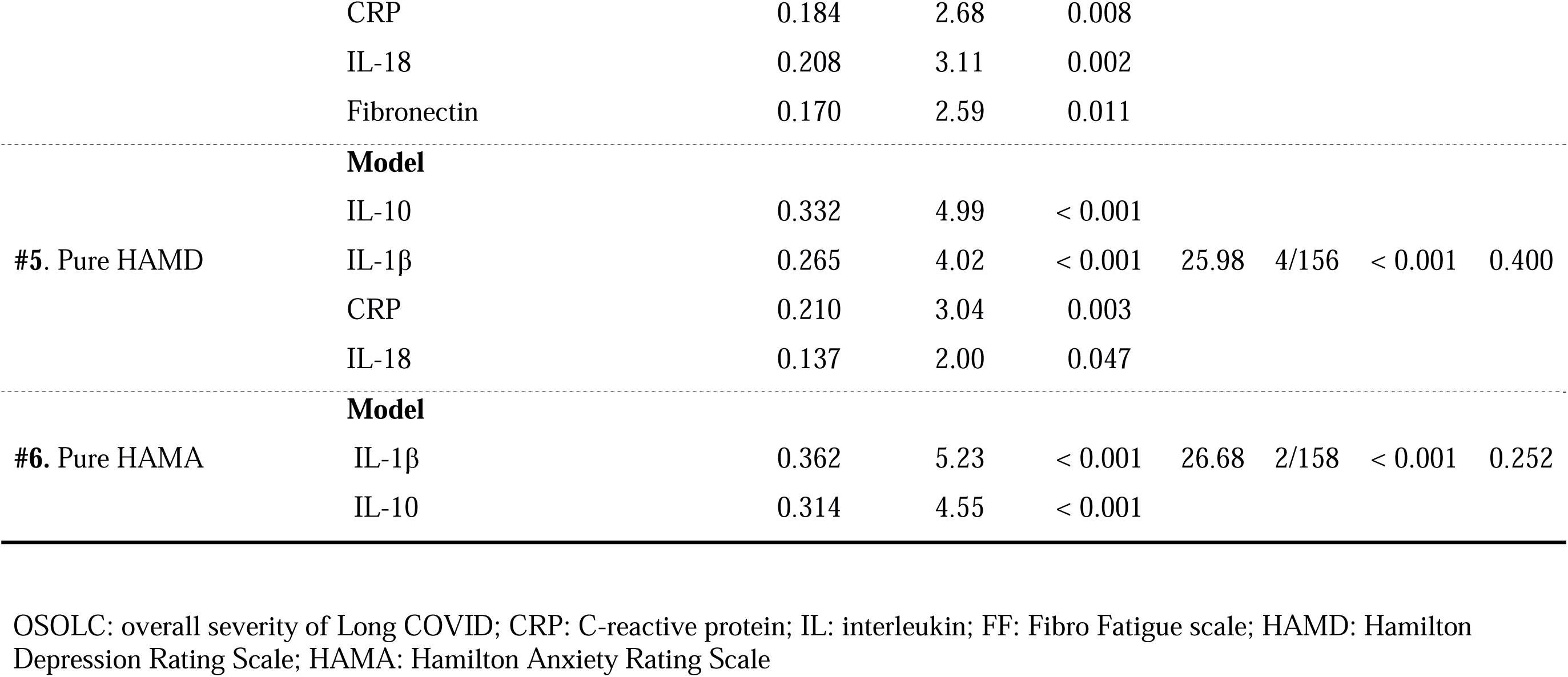
Results of multiple regression analyses with clinical data as dependent variables and biomarkers of Long COVID as explanatory variables.

To explore the relationships between inflammation in the Long COVID period and the assessments of acute infection-associated IPS scores, we calculated a composite index of NLRP3-associated inflammation, which includes z NLRP3, z IL-1β, z IL-18, z caspase-1, z GSDMD, and z fibronectin. In **Table 5**, regression analysis #1 indicates that SpO_2_ levels account for 24.0% of the variance in this composite, demonstrating an inverse association. **Figure 3** illustrates the partial regression of the NLRP3-associated inflammation composite score in relation to the SpO_2_ levels observed during the acute phase of infection. Consequently, diminished SpO_2_ levels observed during the acute phase of the illness serve as predictors for increased NLRP3 inflammasome activity in the subsequent months. An inverse relationship was observed between decreased SpO_2_ values and elevated IL-10 levels, albeit with a modest effect size (specifically: R^2^ = 0.04).

**Figure 3:**
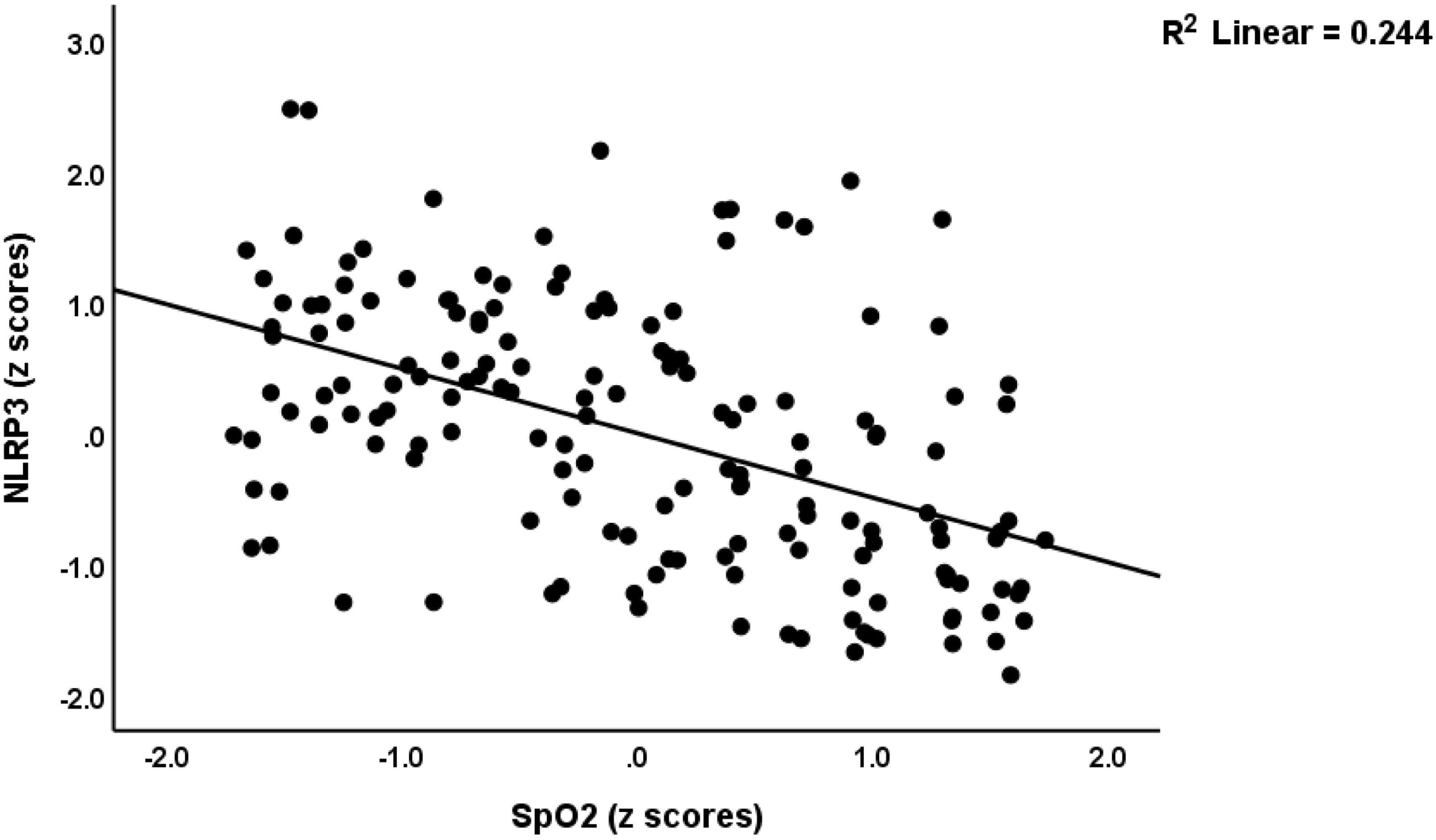
Partial regression of serum NLRP3 inflammasome-associated inflammation during Long COVID on peripheral blood oxygen saturation (SpO_2_) levels during the acute phase of infection (*P* < 0.001).

**Table 5.** Results of multiple regression analyses with total NLRP3 composite and IL-10 as dependent variables and oxygen saturation (SpO_2_).

| Regression | Explanatory variables | $\beta$ | t | P | F<sub>model</sub> | df | P | R<sup>2</sup> |
| --- | --- | --- | --- | --- | --- | --- | --- | --- |
| #1. Composite NLRP 3 | **Model** |  |  |  |  |  |  |  |
|  | SpO<sub>2</sub> | -0.490 | -7.09 | < 0.001 | 50.29 | 1/159 | < 0.001 | 0.240 |
| #5. IL-10 | **Model** |  |  |  |  |  |  |  |
|  | SpO<sub>2</sub> | -0.210 | -2.71 | 0.008 | 7.32 | 1/159 | 0.008 | 0.044 |

NLRP3: nucleotide-binding domain leucine-rich repeat and pyrin domain-containing receptor 3

## Discussion

### Increased IRS biomarkers in Long COVID-19 subgroups

This investigation’s primary conclusion is that we observed markedly different levels of IRS biomarkers of Long COVID among patients who previously experienced severe versus mild acute COVID-19 infection. The differentiation of severe and mild acute COVID-19 infections was predicated on a critical parameter of Long COVID infection, specifically z PBT - z SpO_2_, which is directly correlated with the degree of IRS activation during the acute phase of SARS-CoV-2 infection (Al-Hadrawi *et al*., 2023). We defined the lower values (< 0.41) as the mild acute COVID-19 group, and higher scores (> 0.41) as the severe acute COVID-19 group.

In a previous study, some authors found significantly higher levels of IL-18, caspase-1, and IL-1β in Long COVID, suggesting an activated NLRP3 inflammasome (Al-Hakeim *et al*., 2023). The concentrations of the NLRP3 inflammasome, IL-1β, IL-18, and caspase-1 are markedly elevated in the severe acute COVID-19 cohort compared to the mild acute COVID-19 cohort in the present study. These results align with Rodrigues’ finding that the inflammasome is robustly activated in hospitalized COVID-19 patients (Rodrigues *et al*., 2021a).

Fibronectin, serving as a fundamental element of the extracellular matrix (ECM), plays a vital role in its interactions with angiotensin-converting enzyme (ACE)2, which functions as the receptor for SARS-CoV-2 (Xu *et al*., 2020). Fibroblasts can be stimulated by inflammation and endothelial activation, leading to increased fibronectin synthesis(Enzerink & Vaheri, 2011). High fibronectin levels might indicate that fibrosis-like changes may cause a remodeling of the ECM, whereby the ECM might be rebuilt in a less organized way(Ito *et al*., 2019). Prior research has indicated a correlation between plasma fibronectin levels and the severity of clinical outcomes observed in patients afflicted with COVID-19 (Lemańska-Perek *et al*., 2022).

When DAMPs initiate the cellular pathway, activated NLRP3 catalyzes procaspase-1, turning procaspase-1 into the active form, and the latter cleaves pro-IL-18 and pro-IL-1β into their active forms(Rodrigues & Zamboni, 2023). Furthermore, the cleavage of GSDMD by caspase-1 facilitates the release of IL-18 and IL-1β (He *et al*., 2024). GSDMD is a pore-forming protein that might induce pyroptosis, a type of inflammation-associated cell death (He *et al*., 2024). Previous reports showed that SARS-CoV-2 infection in monocytes leads to pyroptosis via the activation of NLRP3, caspase-1, and GSDMD (Junqueira *et al*., 2022).

In this study, there was a trend towards higher IL-10 concentration in the severe acute COVID-19 group than in the mild COVID-19 group. IL-10 functions as a negative immunoregulatory cytokine, which inhibits NLRP3 activation and reduces IL-1β/IL-18 secretion, balancing the pro-inflammatory signals to limit excessive tissue damage(Lu *et al*., 2021).

In summary, our results show that there might be interconnected aberrations in the fibronectin/NLRP3/caspase-1/IL-1β/IL-18/GSDMD network in the process of Long COVID.

### Associations between the severity of illness and Long COVID biomarkers

The second significant finding indicates that the biomarkers associated with the NLRP3 inflammasome exhibit a robust correlation with the severity of Long COVID. Furthermore, the z PBT–z SpO_2_ index, evaluated during the acute phase of COVID-19, exhibited a robust correlation with inflammatory biomarkers measured several months thereafter. These biomarkers include NLRP3, CRP, Caspase-1, GSDMD, IL-1β, IL-18, and IL-10, which are associated with an escalation in the severity of Long COVID. The latter was conceptualized as a factor derived from pure HAMD, pure HAMA (both affective), and pure FF (physio-somatic) scores. Importantly, we found that all the inflammatory indicators measured in this study, NLRP3, CRP, Caspase-1, GSDMD, Fibronectin, IL-1β, IL-18, and IL-10, are significantly correlated with the overall severity of the physio-affective phenome. Furthermore, 45.7% of the variance in this overall severity index was explained by the cumulative effects of IL-1β, IL-10, CRP, IL-18, and fibronectin (all positively associated). The findings elucidate the significant role of the NLRP3 inflammasome in the initiation or persistence of Long COVID.

Overall, our findings align with previous studies on IRS pathways in Long COVID (Ong *et al*., 2021; Sonnweber *et al*., 2021; Ceban *et al*., 2022). Potere et al. found that several weeks after acute SARS-CoV-2 infection, persistent activation of the NLRP3 pathway can still be detected in peripheral macrophages of recovered patients, suggesting it may underlie the low-grade chronic inflammation observed in Long COVID (Potere *et al*., 2023). It is widely acknowledged that NLRP3-mediated caspase-1 activation drives IL-1β/IL-18 maturation and GSDMD-dependent pyroptosis, and serum levels of IL-18 and GSDMD in patients correlate with disease severity and persistent symptoms (Yin *et al*., 2023). The central P2X7-NLRP3 axis, activated by SARS-CoV-2, promotes neuroinflammation, mitochondrial dysfunction, and neuronal apoptosis, and is implicated in the pathogenesis of Long COVID-associated depression, anxiety, and cognitive impairment(Zhao *et al*., 2021). NLRP3-induced IL-1β triggers excessive neutrophil extracellular trap (NET) formation, leading to microvascular coagulation and alveolar injury, which may underlie persistent dyspnea and hypoxemia in Long COVID (Zhao *et al*., 2021). Furthermore, individuals with pre-existing conditions, including diabetes or obesity, demonstrate heightened baseline NLRP3 activity, which predisposes them to systemic inflammation and multiorgan involvement in Long COVID. This observation underscores the intricate relationship between metabolic processes and inflammatory responses (Potere *et al*., 2023).

In certain studies, higher levels of NLRP3 inflammasome are associated with more severe Long COVID symptoms. Maes et al. discovered that genetic NLRP3 variants are connected to symptoms like fatigue, hyperalgesia, myalgia, and malaise during the acute stage of COVID-19 at the genetic level (Maes *et al*., 2022). Rodrigues et al. investigated 124 COVID-19 convalescents and discovered that persistent Long COVID symptoms, such as fatigue, dyspnea, and cognitive impairment, were associated with more severe clinical scores when peripheral NLRP3 and its downstream cytokines (IL-1β, IL-18) were elevated (Rodrigues *et al*., 2021b). Among patients who experienced post-exertional dyspnea or chest pain three months after discharge, a significant positive correlation was observed between CT-based pulmonary fibrosis scores and peripheral NLRP3 expression levels, indicating the respiratory and cardiovascular sequelae (Babazadeh, 2023). Excessive NLRP3 activation was associated with acute-phase disease severity and predicted persistent multiorgan dysfunction six months post-discharge (Babazadeh, 2023). In summary, the higher the inflammasome activity, the longer and severe the multisystem symptoms of Long COVID persist.

The NLRP3 inflammasome is considered a possible target for therapy due to its pivotal role in the persistent IRS activation and respiratory-neurological-cardiovascular symptoms associated with Long COVID. Several inhibitors have already been advanced into clinical trials. MCC950, a selective NLRP3 binding pocket inhibitor, is currently in the preclinical to phase 1 clinical trial stage. Its primary efficacy, demonstrated in animal studies, includes a significant reduction in IL-1β level and improved pulmonary function (Bakhshi & Shamsi, 2022). Dapansutrile (OLT1177), an oral inhibitor, has completed phase 2 clinical trials. It enhances ejection fraction and exercise tolerance in patients with heart failure (Klück *et al*., 2020). Melatonin inhibits ROS-mediated NLRP3 activation and has progressed to phase 2 clinical trials. It reduces inflammatory marker levels in COVID-19 patients and is particularly suitable for Long COVID patients presenting with sleep disturbances and mood-related symptoms (Zhao *et al*., 2021). Colchicine, an inhibitor of microtubule polymerization and caspase-1 assembly, is undergoing phase 3 clinical trials. It shortens oxygen therapy duration and hospital stays in hospitalized patients, demonstrating potential for Long COVID applications. Baricitinib, a JAK1/2 inhibitor that indirectly blocks NLRP3 transcription, is in phase 3 clinical trials. It significantly reduces COVID-19 mortality, and clinical trials targeting Long COVID are currently being designed (Zhao *et al*., 2021). Current expert consensus identifies the “NLRP3-IL-1 axis” as a priority target for precision therapy in Long COVID and advocates for randomized controlled trials dedicated explicitly to Long COVID.

## Limitations

This paper has some limitations. To begin with, this is a case-control study, so definitive causal links between the IRS pathways and the emergence of Long COVID symptoms cannot be confirmed. Secondly, considering that this research was carried out in Iraq, it is imperative that it be replicated in various other nations.

## Conclusions

Notable distinctions exist in the biomarkers associated with the NLRP3 inflammasome, such as NLRP3, Caspase-1, GSDMD, fibronectin, IL-1β, and IL-18, when comparing Long COVID patients who experienced severe acute COVID-19 infection to those who had mild cases. The identified biomarkers demonstrate a noteworthy correlation with the severity of COVID-19, applicable to both the acute and chronic stages of the disease. Reduced SpO_2_ levels observed during the acute phase of illness serve as a predictor for increased NLRP3 inflammasome activity, which correlates with the emergence of physio-affective symptoms in subsequent months. The findings indicate that the strategic targeting of the NLRP3 inflammasome may be beneficial in addressing both the acute infectious phase of COVID-19 as well as the prolonged symptoms associated with Long COVID. Novel pharmacological agents are currently under development aimed at specifically targeting the activation of NLRP3.

## Acknowledgements

None

## Ethical approval and consent to participate

Every participant signed a written informed consent form. The research obtained authorization from the Najaf Health Directorate Training and Human Development Center and the institutional ethics board of the University of Kufa, as shown by documents with the numbers 4316-2024/1/29. The research complied with Iraqi and international ethical and privacy regulations, including the International Conference on Harmonization of Good Practice, the Belmont Report, the CIOMS Guidelines, and the Declaration of Helsinki by the World Medical Association. Furthermore, our institutional review board adheres to the International Guidelines for Human Research Safety (ICH-GCP).

## Author Contributions

HA-H and MM conducted the design of the study. HA-J collected blood samples. HA-H and HA-J performed the analyses. MM conducted the statistical analysis. YZ wrote the first draft, which was revised by MM and all other authors. All the authors took part in editing and have given their approval for the final version to be submitted.

## Data Availability Statement

The corresponding author (MM) is available to provide the study data upon request.

